# Characterizing the limitations of using diagnosis codes in the context of machine learning for healthcare

**DOI:** 10.1101/2023.03.14.23287202

**Authors:** Lin Lawrence Guo, Keith E. Morse, Catherine Aftandilian, Ethan Steinberg, Jason Fries, Jose Posada, Scott Lanyon Fleming, Joshua Lemmon, Karim Jessa, Nigam Shah, Lillian Sung

**Affiliations:** Program in Child Health Evaluative Sciences, The Hospital for Sick Children, Toronto, ON; Division of Pediatric Hospital Medicine, Department of Pediatrics, Stanford University, Palo Alto, CA; Division of Hematology/Oncology, Department of Pediatrics, Stanford University, Palo Alto, CA; Stanford Center for Biomedical Informatics Research, Stanford University, Palo Alto, CA; Universidad del Norte, Columbia; Information Services, The Hospital for Sick Children, Toronto ON; Division of Haematology/Oncology, The Hospital for Sick Children, Toronto, ON

**Keywords:** electronic health records, diagnostic coding practice, cohort identification, outcome identification, machine learning for health

## Abstract

**Importance:** Diagnostic codes are commonly used as inputs for clinical prediction models, to create labels for prediction tasks, and to identify cohorts for multicenter network studies. However, the coverage rates of diagnostic codes and their variability across institutions are underexplored.

**Objective:** Primary objective was to describe lab- and diagnosis-based labels for 7 selected outcomes at three institutions. Secondary objectives were to describe agreement, sensitivity, and specificity of diagnosis-based labels against lab-based labels.

**Methods:** This study included three cohorts: SickKids_Peds_ from The Hospital for Sick Children, and Stanford_Peds_ and Stanford_Adults_ from Stanford Medicine. We included seven clinical outcomes with lab-based definitions: acute kidney injury, hyperkalemia, hypoglycemia, hyponatremia, anemia, neutropenia and thrombocytopenia. For each outcome, we created four lab-based labels (abnormal, mild, moderate and severe) based on test result and one diagnosis-based label. Proportion of admissions with a positive label were presented for each outcome stratified by cohort. Using lab-based labels as the gold standard, agreement using Cohen’s Kappa, sensitivity and specificity were calculated for each lab-based severity level.

**Results:** The number of admissions included were: SickKids_Peds_ (n=59,298), Stanford_Peds_ (n=24,639) and Stanford_Adults_ (n=159,985). The proportion of admissions with a positive diagnosis-based label was significantly higher for Stanford_Peds_ compared to SickKids_Peds_ across all outcomes, with odds ratio (99.9% confidence interval) for abnormal diagnosis-based label ranging from 2.2 (1.7-2.7) for neutropenia to 18.4 (10.1-33.4) for hyperkalemia. Lab-based labels were more similar by institution. When using lab-based labels as the gold standard, Cohen’s Kappa and sensitivity were lower at SickKids_Peds_ for all severity levels compared to Stanford_Peds_.

**Conclusions:** Across multiple outcomes, diagnosis codes were consistently different between the two pediatric institutions. This difference was not explained by differences in test results. These results may have implications for machine learning model development and deployment.

## INTRODUCTION

Machine learning models based on electronic health records (EHRs) are increasingly being developed and implemented into routine care. They have improved outcomes related to reducing acute care visits among ambulatory cancer patients^1^, decreasing in-hospital clinical deterioration^2^, increasing serious illness conversations^3^, improving platelet utilization^4^ and refining antibiotic choice^5^ as examples.

To develop models, inputs or features are extracted from EHRs; these reflect different aspects of care such as diagnostic codes, laboratory tests, microbiology results, medication administrations, blood product administration, and procedures. Diagnostic codes are also frequently used to define the outcome of interest or label. How well each institution generates accurate diagnostic codes may vary depending on the coding process specific to the instution^6^ and clinical diagnostic practice specific to the hospital unit or physician^6-8^. This variability might influence the performance and generalizability of machine learning models developed at institutions with different diagnostic coverage rates. In pediatric populations, the coverage rates of diagnostic codes and their variability across institutions are underexplored^9,10^.

A challenge to studying the question of diagnostic code coverage is the creation of gold standard labels as the diagnostic codes themselves are often used to develop these labels. One type of clinical data in which the label is inherent within the result itself is laboratory-based outcomes. Abnormal lab tests can be defined using institution-specific reference ranges. In addition, levels of severity (mild, moderate, and severe) for each abnormal lab test can be defined based upon widely accepted thresholds. Thus, evaluating diagnostic code coverage against lab-based definitions provides a pragmatic setting in which to evaluate this question.

Consequently, the primary objective was to describe lab- and diagnosis-based labels for selected outcomes at three institutions. Secondary objectives were to describe agreement, sensitivity, and specificity of diagnosis-based labels against lab-based labels.

## METHODS

### Design

This study used data derived from EHRs at three institutions, namely The Hospital for Sick Children (SickKids) in Toronto, Ontario; Lucile Packard Children’s Hospital (primarily pediatric-directed care) in Palo Alto, California and Stanford Health Care (primarily adult-directed care) in Palo Alto, California. The overall goal was to compare lab- and diagnosis-based labels for pediatric patients at SickKids vs. Stanford. We included a Stanford adult cohort for descriptive purposes.

### Data Sources

#### SEDAR

The data source at SickKids was the SickKids Enterprise-wide Data in Azure Repository (SEDAR)^11^. SEDAR contains a curated version of Epic Clarity data that is being used for operational, quality improvement and research purposes. This study was approved as a quality improvement project at SickKids and consequently, the requirement for Research Ethics Board approval and informed consent were not required.

#### STARR

The Stanford medicine research data repository (STARR)^12^ is the clinical data warehouse that contains records routinely collected in the EHR of Stanford Medicine, which is comprised of Lucile Packard Children’s Hospital and Stanford Health Care. The data have been mapped to the standard concept identifiers and structure of the Observational Medical Outcomes Partnership Common Data Model (OMOP-CDM)^13^, resulting in a dataset named STARR-OMOP. This study used a de-identified version of STARR-OMOP^12^ in which protected health information has been redacted. Because of de-identification, requirement for Institutional Review Board approval and informed consent were not required for data use.

### Cohorts

We defined three cohorts. SickKids_Peds_ was obtained using SEDAR while Stanford_Peds_ and Stanford_Adults_ were obtained using STARR-OMOP and applying age-specific restrictions. Table 1 summarizes the inclusion criteria for each cohort. Across all three cohorts, inpatient admissions were included if they occurred between 2018-06-02 to 2022-08-01. The pediatric cohorts (SickKids_Peds_ and Stanford_Peds_) included patients who were 28 days or older and younger than 18 years on the day of admission. We excluded neonates 1 to 27 days of age because Lucile Packard Children’s Hospital has an obstetrical unit and consequently includes healthy newborns while SickKids does not have an obstetrical unit and does not routinely see healthy newborns. Stanford_Adults_ included adult patients aged 18 or above on the day of admission. Multiple admissions per patient were permitted as long as eligibility criteria were met.

**Table 1.** Inclusion criteria and cohort characteristics.

|  | SickKids<sub>Peds</sub> | Stanford<sub>Peds</sub> | Stanford<sub>Adults</sub> |
| --- | --- | --- | --- |
| **Inclusion Criteria** |  |  |  |
| Age at Admission | ≥ 28 days and<br>< 18 years | ≥ 28 days and<br>< 18 years | ≥ 18 years |
| Admission Date | 2018-06-02 to<br>2022-08-01 | 2018-06-02 to<br>2022-08-01 | 2018-06-02 to<br>2022-08-01 |
| **Cohort Characteristics** |  |  |  |
| Number Admissions | 59,298 | 24,639 | 159,985 |
| Number Patients | 36,585 | 14,518 | 103,170 |
| Median Age at Admission [IQR] | 6 [2 - 12] | 6 [2 - 12] | 57 [36 - 71] |
| Pediatric Age Group, n (%) |  |  |  |
| Infant (28 days – 12 months) | 8980 (15.1%) | 3869 (15.7%) |  |
| Toddler (13 months – 2 years) | 5661 (9.5%) | 2269 (9.2%) |  |
| Early childhood (2 – 5 years) | 10263 (17.3%) | 4307 (17.5%) |  |
| Middle childhood (6 – 11 years) | 14837 (25.0%) | 6056 (24.6%) |  |
| Early adolescence (12 – 17 years) | 19557 (33.0%) | 8138 (33.0%) |  |
| Sex, n (%) |  |  |  |
| Females | 27,264<br>(46.0%) | 11,800<br>(47.9%) | 91,770 (57.4%) |
| Males | 32,030<br>(54.0%) | 12,837<br>(52.1%) | 68,207 (42.6%) |
| Unknown | 4 (<0.1%) | 2 (<0.1%) | 15 (<0.1%) |
| Patient Outcomes |  |  |  |
| In-hospital mortality, n (%) | 297 (0.5%) | 203 (0.8%) | 3088 (1.9%) |
| Median length of stay [IQR] | 2 [1-5] | 3 [1-6] | 3 [2-6] |

Abbreviations: SickKids – The Hospital for Sick Children; Peds – pediatrics; IQR – interquartile range

### Outcome Definitions

We included seven clinical outcomes that have lab-based definitions, namely acute kidney injury (AKI), hyperkalemia, hypoglycemia, hyponatremia, anemia, neutropenia, and thrombocytopenia. We appreciate there are a large number of potential lab-based outcomes; these seven were chosen based on our current research interests and because they are clinically meaningful. The outcomes were chosen *a priori*, before conducting any of the analyses. We purposely did not include abnormal high and low for the same lab test (for example, hyperglycemia and hypoglycemia) as they may be correlated. For each outcome, we created four lab-based labels based on the test result and one diagnosis-based label; these five labels were evaluated in each patient admission. Appendix 1 shows the thresholds for each severity level (mild, moderate, and severe levels) of the lab-based labels; these thresholds were based upon research studies or guidelines^14-20^. We also labeled the result as abnormal if the result was above or below (not both) of the institution-specific reference range. For lab-based labels, units for lab results were normalized, and severity level was nested. For example, a patient admission with severe hypoglycemia would also be included in the analyses for mild and moderate hypoglycemia. For the diagnosis-based label, we considered an outcome to be present if at least one outcome-related diagnosis code was assigned to the admission.

### Concept Selection for Lab-based and Diagnosis-based Labels

We adopted different search strategies for concepts in STARR-OMOP and SEDAR due to differences in structure and vocabularies for clinical codes. Diagnosis codes were derived from the “condition_occurrence” table for STARR-OMOP and from the “diagnosis” table for SEDAR. Lab test results were obtained from the “measurement” table for STARR-OMOP and from the “lab” table for SEDAR. For face validation, diagnosis codes and lab result distributions obtained from STARR-OMOP were reviewed by three clinicians (KEM, CA and LS) to identify errors related to normalization or concept selection. At SickKids, this same review was only conducted by one clinician (LS) due to access restrictions.

### Baseline Characteristics by Cohort

To explore whether there were differences in the cohorts with respect to patients, we described the demographic characteristics and raw lab results of patients between centers. Demographic characteristics included age, sex, length of stay, and the prevalence of in-hospital mortality. For the evaluation of raw lab results, we determined the minimum or maximum result for each lab test per admission and stratified by cohort.

To gain insight into whether there were differences between pediatric institutions with respect to laboratory procedures or clinical practice, we described the institution- and age group-specific reference ranges for abnormal lab results by SickKids_Peds_ and Stanford_Peds_. The pediatric age groups were defined by the National Institute of Child Health and Human Development^21^ as infancy (28 days – 12 months), toddler (13 months – 2 years), early childhood (2 – 5 years), middle childhood (6 – 11 years) and early adolescence (12 – 17 years). In addition, we evaluated lab testing frequency calculated as the number of tests per inpatient day for each admission.

### Statistical Analysis

The primary objective was to describe lab- and diagnosis-based labels at the three institutions. These were presented as the proportion of admissions with at least one positive label. To describe the odds of a lab- or diagnosis-based label by whether the pediatric admission occurred at Stanford_Peds_ vs. SickKids_Peds_, analysis was complicated by the large number of admissions and multiple testing (35 separate evaluations for this analysis alone). In addition, there were multiple admissions per patient, resulting in correlation within individuals. To address these concerns, we took several steps. First, we focused on describing the odds ratio (OR) and 99.9% confidence interval (CI) for a lab- or diagnosis-based label by pediatric institution. Second, we described the 99.9% confidence interval rather than the 95% confidence interval to help address multiple testing. Third, we did not calculate P values but rather, focused on describing CIs with the exception of comparing lab testing frequency by institution. Finally, to address multiple admissions per patient, OR and 99.9% CI were calculated using mixed-effects logistic regression. Models included each binary label as the outcome, institution and pediatric age group as fixed effects and subject as random intercept. Analysis was performed using the glmer function from lme4 package in R.

The secondary objectives were to describe agreement, sensitivity, and specificity of diagnosis-based labels against lab-based labels. Agreement in each cohort was described using Cohen’s Kappa coefficient. Sensitivity and specificity of the diagnosis-based labels were determined using each of the lab-based labels as the gold standard. For each metric, we presented the median and ranges stratified by cohort and lab-based severity (abnormal, mild, moderate and severe).

As an exploratory analysis, we separately evaluated each visited unit during admissions at each pediatric institution. We examined the weighted proportion of positive lab-based labels and positive diagnosis-based labels for each hospital unit and calculated Spearman’s rho (r) based on the average across lab-based severity.

To describe lab-based reference ranges for pediatric patients, we described the threshold for an abnormal lab test by pediatric age group stratified by institution. Where the threshold varied within an age group, the range was visually depicted using a bar rather than a line. To compare testing frequency between pediatric institutions, mixed-effects linear regression was performed with number of lab tests per admission as the outcome, institution and pediatric age group as fixed effects and subject as random intercept. Analysis was performed using the lmer function from the lme4 package in R.

All analyses were conducted using Python (version 3.7) and R (version 4.1.2).

## RESULTS

### Baseline Characteristics

The number of admissions included were: SickKids_Peds_ (n=59,298), Stanford_Peds_ (n=24,639) and Stanford_Adults_ (n=159,985). Characteristics of the three cohorts are listed in Table 1. The distributions of age, sex, in-hospital mortality, and median length of stay were similar between SickKids_Peds_ and Stanford_Peds_ while the distribution of sex and in-hospital mortality differed at Stanford_Adult_. Table 2 shows the distribution of minimum or maximum results for each lab test per admission by cohort. Distributions appeared similar between Stanford_Peds_ and SickKids_Peds_ with the exception of minimum absolute neutrophil count, which was lower at SickKids_Peds_ vs. Stanford_Peds_. Appendix 2 shows that the reference ranges varied between SickKids_Peds_ and Stanford_Peds._ Reference ranges for glucose and sodium were the same for all age groups except infants. Reference ranges for potassium and platelets were notably different by institution across age groups. Appendix 3 shows the average number of lab tests performed per inpatient day across all admissions stratified by institution. SickKids_Peds_ performed significantly fewer tests compared to Stanford_Peds_ for all tests.

**Table 2.** Distribution of minimum or maximum results for each lab test per admission and stratified by cohort.

| Lab Test | Units | SickKids<sup>Peds</sup><br>Median (IQR) | Stanford<sup>Peds</sup><br>Median (IQR) | Stanford<sup>Adults</sup><br>Median (IQR) |
| --- | --- | --- | --- | --- |
| Maximum Creatinine | umol/L | 42.0<br>(29.0 - 60.0) | 34.5<br>(23.0 - 53.0) | 78.7<br>(61.0 - 109.6) |
| Maximum Potassium | mmol/L | 4.5<br>(4.1 – 5.0) | 4.4<br>(4.1 – 4.9) | 4.4<br>(4.1 – 4.8) |
| Minimum Glucose | mmol/L | 4.9<br>(4.3 - 5.5) | 4.9<br>(4.3 - 5.6) | 5.2<br>(4.6 - 5.9) |
| Minimum Sodium | mmol/L | 138.0<br>(136.0 – 140.0) | 137.0<br>(134.0 – 139.0) | 135.0<br>(132.0 – 138.0) |
| Minimum Absolute Neutrophil Count | 10<sup>9</sup>/L | 2.2<br>(1.1 – 3.9) | 3.4<br>(1.9 – 5.9) | 5.5<br>(3.6 – 7.7) |
| Minimum Hemoglobin | g/L | 102.0<br>(85.0 – 119.0) | 99.0<br>(81.0 - 117.0) | 103.0<br>(85.0 - 119.0) |
| Minimum Platelet Count | 10<sup>9</sup>/L | 231.0<br>(147.0 - 321.0) | 219.0<br>(140.0 - 303.0) | 188.0<br>(139.0 - 243.0) |

Abbreviation: SickKids: The Hospital for Sick Children; Peds: pediatrics; IQR: interquartile range.

### Prevalence of Lab-based and Diagnosis-based Labels

Table 3 provides the percentage of admissions with a positive lab- and diagnosis-based label. Table 3 and Figure 1 show OR and 99.9% CI. The proportion of admissions with a positive diagnosis-based label was significantly higher for Stanford_Peds_ compared to SickKids_Peds_ across all outcomes, with OR (99.9% CI) for abnormal diagnosis-based label ranging from 2.2 (1.7-2.7) for neutropenia to 18.4 (10.1-33.4) for hyperkalemia. Lab-based labels were more similar by institution although several were significantly different as demonstrated by CIs that did not cross 1.

**Table 3.** Proportion of admissions with positive lab- and diagnosis-based labels by cohort.

| Severity* | Outcome** | SickKids<sub>Peds</sub> | Stanford<sub>Peds</sub> | Stanford<sub>Adults</sub> | Odds Ratio (99.9% CI)*** |
| --- | --- | --- | --- | --- | --- |
| Number Admissions |  | 59,298 | 24,639 | 159,985 |  |
| Lab (Abnormal) | AKI | 4,553 (7.7%) | 2,266 (9.2%) | 41,761 (26.1%) | 1.3 (1.1,1.4) |
|  | Hyperkalemia | 5,475 (9.2%) | 1,596 (6.5%) | 9,790 (6.1%) | 0.7 (0.6,0.8) |
|  | Hypoglycemia | 3,006 (5.1%) | 1,613 (6.5%) | 12,161 (7.6%) | 1.4 (1.2,1.5) |
|  | Hyponatremia | 3,888 (6.6%) | 4,141 (16.8%) | 53,512 (33.4%) | 2.9 (2.6,3.2) |
|  | Neutropenia | 4,263 (7.2%) | 1,804 (7.3%) | 5,085 (3.2%) | 1.0 (0.9,1.2) |
|  | Anemia | 14,839 (25.0%) | 10,496 (42.6%) | 119,796 (74.9%) | 2.4 (2.2,2.6) |
|  | Thrombocytopenia | 10,667 (18.0%) | 3,844 (15.6%) | 44,636 (27.9%) | 0.8 (0.7,0.9) |
| Lab (Mild) | AKI | 4,478 (7.6%) | 3,461 (14.0%) | 31,930 (20.0%) | 1.8 (1.6,2.1) |
|  | Hyperkalemia | 3,408 (5.7%) | 1,848 (7.5%) | 9,776 (6.1%) | 1.3 (1.2,1.5) |
|  | Hypoglycemia | 3,844 (6.5%) | 2,197 (8.9%) | 13,115 (8.2%) | 1.5 (1.3,1.6) |
|  | Hyponatremia | 6,169 (10.4%) | 5,648 (22.9%) | 66,559 (41.6%) | 2.6 (2.4,2.8) |
|  | Neutropenia | 4,868 (8.2%) | 1,921 (7.8%) | 5,099 (3.2%) | 0.9 (0.8,1.1) |
|  | Anemia | 21,232 (35.8%) | 11,436 (46.4%) | 114,488 (71.6%) | 1.6 (1.5,1.7) |
|  | Thrombocytopenia | 7,061 (11.9%) | 3,844 (15.6%) | 44,636 (27.9%) | 1.4 (1.2,1.5) |
| Lab (Moderate) | AKI | 1,650 (2.8%) | 1,550 (6.3%) | 11,321 (7.1%) | 2.1 (1.8,2.4) |
|  | Hyperkalemia | 1,939 (3.3%) | 1,137 (4.6%) | 4,790 (3.0%) | 1.4 (1.3,1.6) |
|  | Hypoglycemia | 2,084 (3.5%) | 1,178 (4.8%) | 7,366 (4.6%) | 1.4 (1.2,1.6) |
|  | Hyponatremia | 885 (1.5%) | 917 (3.7%) | 15,027 (9.4%) | 2.5 (2.1,3.0) |
|  | Neutropenia | 3,346 (5.6%) | 1,172 (4.8%) | 2,850 (1.8%) | 0.9 (0.7,1.0) |
|  | Anemia | 17,039 (28.7%) | 9,429 (38.3%) | 91,276 (57.1%) | 1.6 (1.4,1.7) |
|  | Thrombocytopenia | 4,175 (7.0%) | 2,349 (9.5%) | 19,327 (12.1%) | 1.4 (1.2,1.7) |
| Lab (Severe) | AKI | 616 (1.0%) | 612 (2.5%) | 5,804 (3.6%) | 2.4 (1.9,2.9) |
|  | Hyperkalemia | 810 (1.4%) | 588 (2.4%) | 1,331 (0.8%) | 1.8 (1.5,2.1) |
|  | Hypoglycemia | 1,088 (1.8%) | 585 (2.4%) | 4,042 (2.5%) | 1.3 (1.1,1.6) |
|  | Hyponatremia | 301 (0.5%) | 269 (1.1%) | 3,790 (2.4%) | 2.1 (1.6,2.8) |
|  | Neutropenia | 2,290 (3.9%) | 635 (2.6%) | 1,316 (0.8%) | 0.7 (0.6,1.0) |
|  | Anemia | 2,675 (4.5%) | 1,912 (7.8%) | 14,239 (8.9%) | 1.8 (1.6,2.0) |
|  | Thrombocytopenia | 2,328 (3.9%) | 1,306 (5.3%) | 7,189 (4.5%) | 1.4 (1.2,1.7) |
| Diagnosis | AKI | 176 (0.3%) | 1,139 (4.6%) | 9,440 (5.9%) | 15.1 (11.3,20.0) |
|  | Hyperkalemia | 38 (0.1%) | 353 (1.4%) | 2,453 (1.5%) | 18.4 (10.1,33.4) |
|  | Hypoglycemia | 209 (0.4%) | 412 (1.7%) | 1,385 (0.9%) | 4.3 (3.1,5.8) |
|  | Hyponatremia | 388 (0.7%) | 708 (2.9%) | 5,219 (3.3%) | 4.2 (3.4,5.2) |
|  | Neutropenia | 776 (1.3%) | 790 (3.2%) | 1,572 (1.0%) | 2.2 (1.7,2.7) |
|  | Anemia | 974 (1.6%) | 4,238 (17.2%) | 30,935 (19.3%) | 9.7 (8.3,11.3) |
|  | Thrombocytopenia | 192 (0.3%) | 2,132 (8.7%) | 10,334 (6.5%) | 14.8 (10.7,20.6) |

\* Abnormal, mild, moderate and severe according to Appendix 1. Abnormal means either above or below (not both) reference range
\*\* Lab-based measure of acute kidney injury was hypercreatinemia
\*\*\* Odds ratio for SickKids<sub>Peds</sub> vs. Stanford<sub>Peds</sub> obtained using mixed-effects logistic regression with each binary label as outcome, institution and pediatric age group as fixed effects, and subject as random intercept.
Abbreviation: AKI: acute kidney injury; SickKids: The Hospital for Sick Children; Peds: pediatrics.

**Figure 1.**
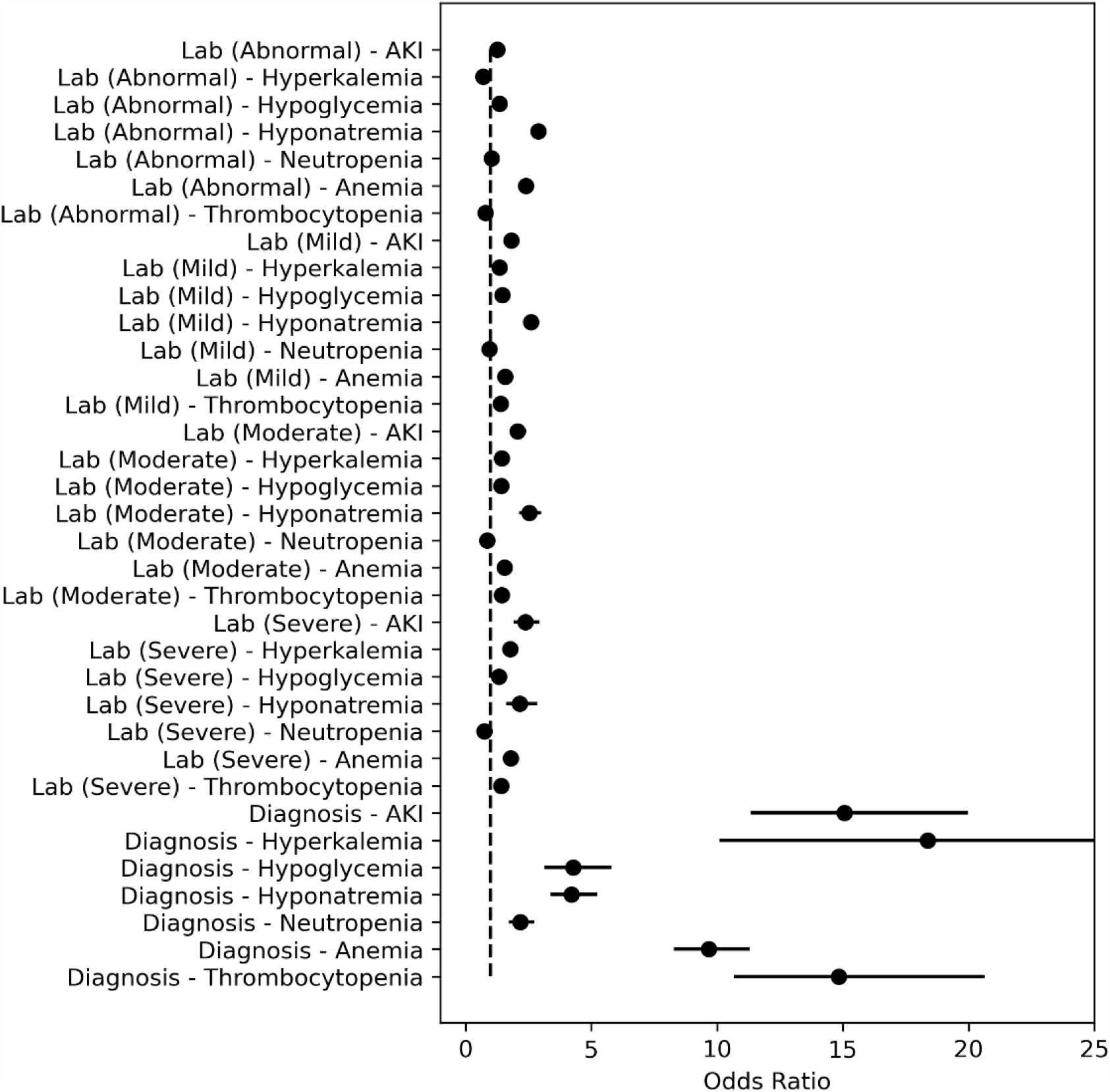
Odds of a lab- or diagnosis-based label by whether the pediatric admission occurred at Stanford vs. SickKids. Figure shows odds ratio and 99.9% confidence interval showing odds of an abnormal label by institution. Dashed line indicates an odds ratio of 1. An odds ratio of >1 corresponds to higher odds of assigning a positive label for Stanford_Peds_ compared to SickKids_Peds_. Odds ratios were obtained using mixed-effects logistic regression with each binary label as outcome, institution and pediatric age group as fixed effects and subject as random intercept

### Agreement between Outcome Definitions

Figure 2 shows the evaluations of diagnosis-based labels against each of the lab-based labels using Cohen’s Kappa coefficient, sensitivity, and specificity. Overall, diagnosis codes had high specificity (mean=0.984, standard deviation (SD)=0.026) but low sensitivity (mean = 0.203, SD=0.158) and low Kappa (mean=0.213, SD=0.132) with lab-based labels. Compared to Stanford_Peds_, SickKids_Peds_ diagnosis-based labels had lower Kappa statistic and sensitivity, but higher specificity.

**Figure 2.**
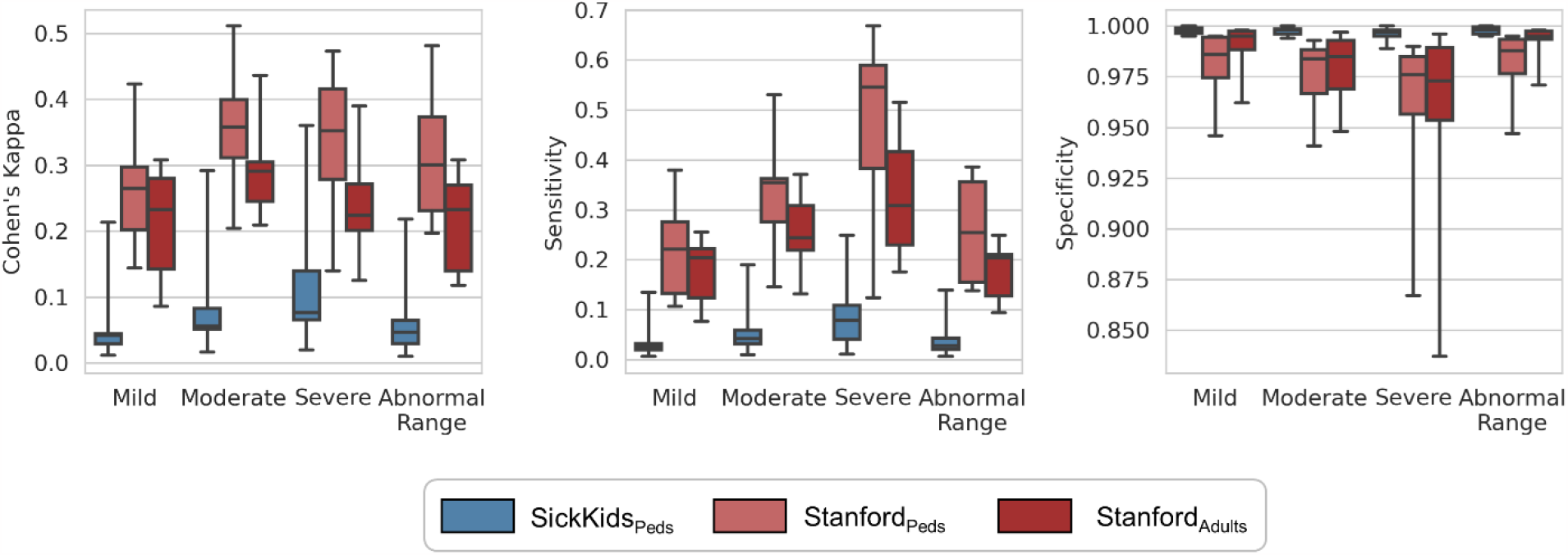
Cohen’s Kappa, sensitivity, and specificity for diagnosis-based labels against lab-based labels. The figure shows median, interquartile range (shaded box) and range (whiskers)

Figure 3 plots the weighted proportions of positive diagnosis-based labels against the weighted proportions of positive lab-based labels for each hospital unit at SickKids_Peds_ and Stanford_Peds_. At Stanford_Peds_, units associated with more patients with lab-based labels also had more patients with a positive diagnosis-based label for a clinical outcome, with Spearman r ranging from 0.513 (hyponatremia) to 0.871 (neutropenia). In contrast, the Spearman r’s were generally lower at SickKids_Peds_ across all outcomes, and ranged from 0.010 (hypoglycemia) to 0.356 (anemia).

**Figure 3.**
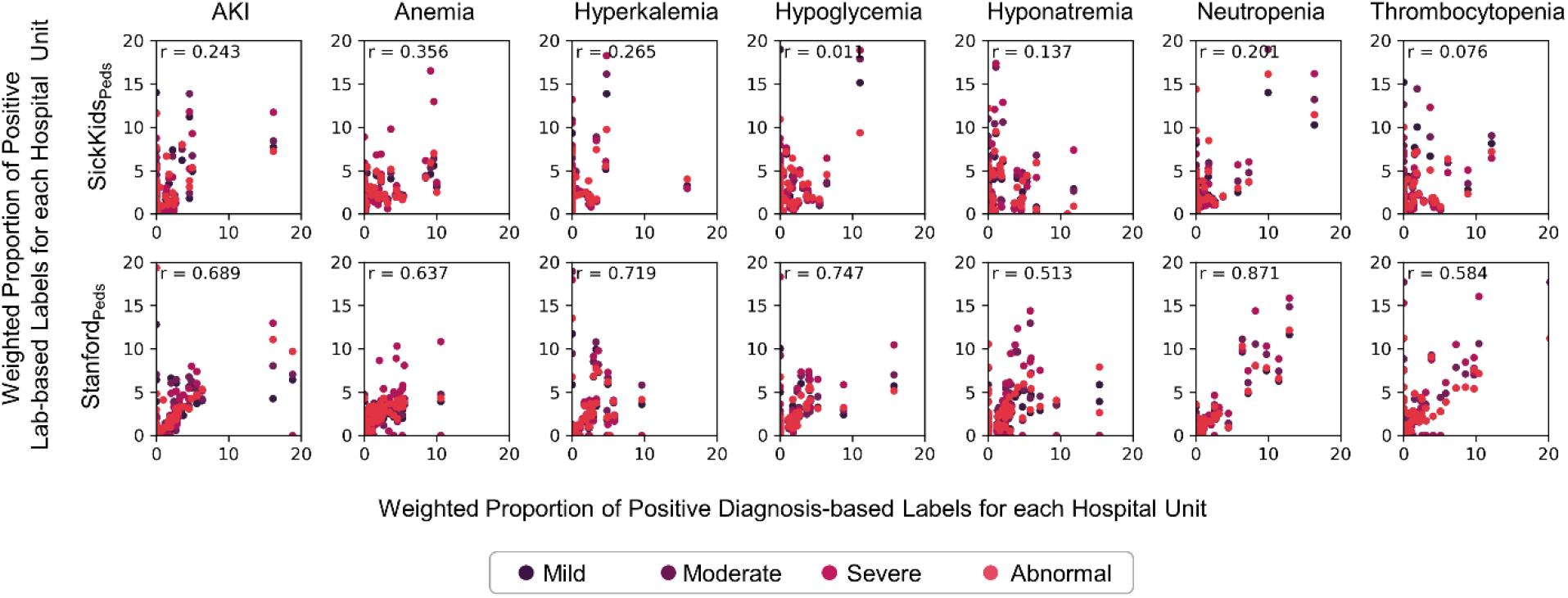
Agreement between diagnosis-based labels and lab-based labels across hospital units. The numbers on the x- and y- axis represent the weighted proportion of positive lab-based labels and positive diagnosis-based labels for each hospital unit visited during the admission. Spearman rho (r) was calculated based on the average across lab-based severity

## DISCUSSION

Our results showed that despite similar demographic characteristics, there were large differences between the two pediatric institutions in the proportion of admissions with diagnosis codes for the evaluated clinical outcomes. In addition, diagnosis-based labels generally had low agreement with lab-based labels and displayed low sensitivity but high specificity when considering lab-based labels as the gold standard, with differences observed between the two institutions. In addition, we found differences between the two institutions in terms of test ordering frequency and even laboratory test references ranges.

These results suggest that if machine learning models are intended for deployment at multiple institutions, reliance on diagnostic codes, either as feature or labels, could be problematic if institutions have different coding practices. Second, they suggest that using institutional reference ranges to categorize laboratory test results may contribute to geographic dataset shift. This study contributes to the body of evidence that demonstrates the limitations of using diagnosis codes for outcome identification. Studies have reported low sensitivity rate when using diagnosis codes to identify, for example, acute kidney injury^22^, obesity^23^, and symptoms of coronavirus disease 2019^24^. In addition, this study showed differences between and within institutions in diagnostic practice that may have contributed to the differences in the performance of diagnosis codes for outcome identification.

Diagnosis codes from the EHR are commonly queried during feature extraction^25-29^, label creation^30^, and cohort identification^31^. Heterogeneity in diagnostic practice across hospital units within the same institution (e.g., SickKids) can impact a model’s performance within sub-populations or spuriously associate certain units with the outcome of interest during model development. In addition, the cross-institution difference in diagnostic coding practice has implications for network studies as it violates the assumption that coding practice is comparable across institutions and creates heterogeneity in outcome prevalence as an artifact of code availability.

While we found that the proportions of positive lab-based labels were more similar between pediatric institutions, there were significant differences although smaller than that observed for diagnosis-based labels. Possible contributions were the observed differences in lab testing frequency between the two pediatric institutions. In addition, the reference ranges themselves were different for tests with the same absolute interpretation regardless of where the test was conducted. For example, two hypothetical children with the same platelet count could be considered to have a normal test at one institution and an abnormal test at the second institution. Some SickKids reference ranges were based upon those established by the Canadian Laboratory Initiative on Pediatric Reference Intervals (CALIPER) initiative,^32^ which contributed to the disparity. Nonetheless, this has implications for machine learning models. First, it is common during feature processing to categorize lab test results as normal, high, and low based upon the reference range^25,33^. Having different reference ranges would thus produce different features despite having the same numerical value. Second, different reference ranges may impact downstream clinical decision making and variability of resultant clinical actions, for example procedures and medication administrations. Since these actions will be recorded in the EHR, impact on clinical decision making can further worsen geographic dataset shift.

The strengths of this study include the ability to evaluate multiple institutions in different countries and the involvement of clinician co-investigators who contributed to the identification of concepts to include in the various label definitions. However, this study is limited for several reasons. First, we only evaluated seven outcomes. In addition, the outcomes were restricted to those that have lab-based definitions in order to use lab tests to develop gold standard labels. Outcomes that are more complex might require chart review to establish gold standards and more sophisticated electronic phenotyping approaches to reach reasonable performance^34,35^. Finally, our analyses were restricted to admissions within a relatively narrow time period (2018-2022). It might be useful to characterize practice differences over time as temporal distribution shift can negatively impact model performance over time^27,36^.

In conclusion, across multiple outcomes, diagnosis codes were consistently different between the two pediatric institutions. This difference was not explained by differences in test results. These results may have implications for machine learning model development and deployment.

## Data Availability

The data are available from the corresponding author upon reasonable request.

## Figure Legend

**Appendix 1.** Thresholds for each severity level of the lab-based labels.

| Outcome | Test | Unit | Mild | Moderate | Severe |
| --- | --- | --- | --- | --- | --- |
| Acute Kidney Injury | Creatinine | μmol/L | ≥1.5x baseline<sup>1</sup> or increase of 26.5 | ≥2x baseline | ≥3x or increase of 353.6 |
| Hyperkalemia | Potassium | mmol/L | >5.5 | >6 | >7 |
| Hypoglycemia | Glucose | mmol/L | <3.9 | <3.5 | <3 |
| Hyponatremia | Sodium | mmol/L | ≤135 | <130 | <125 |
| Neutropenia | Absolute neutrophil count<sup>2</sup> | 10<sup>9</sup>/L | <1.5 | <1.0 | <0.5 |
| Anemia | Hemoglobin | g/L | <120 | <110 | <70 |
| Thrombocytopenia | Platelet | 10<sup>9</sup>/L | <150 | <100 | <50 |

<sup>1</sup> Baseline defined as the minimum creatinine over the 3-month period prior to the index time, or the upper bound of the normal reference range in the absence of an observed value
<sup>2</sup> Absolute neutrophil count was the sum of neutrophil and band counts. If a complete blood count differential was not performed or not reported, we categorized neutropenia if the total white blood cell count met the threshold

**Appendix 2.**
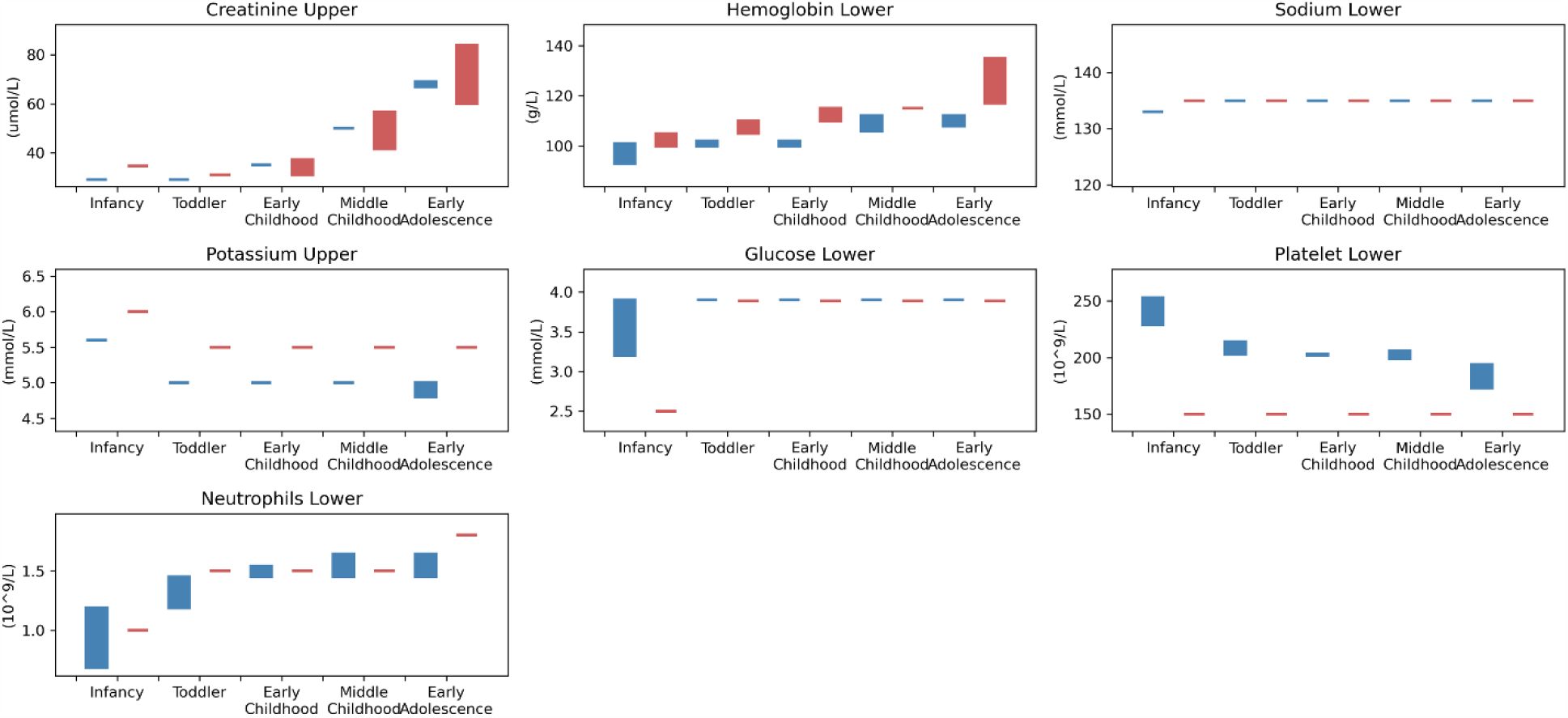
Institution- and age group-specific threshold for abnormal lab test for SickKids_Peds_ (blue) and Stanford_Peds_ (red) Abbreviation: SickKids: The Hospital for Sick Children; Peds: pediatrics.

**Appendix 3.**
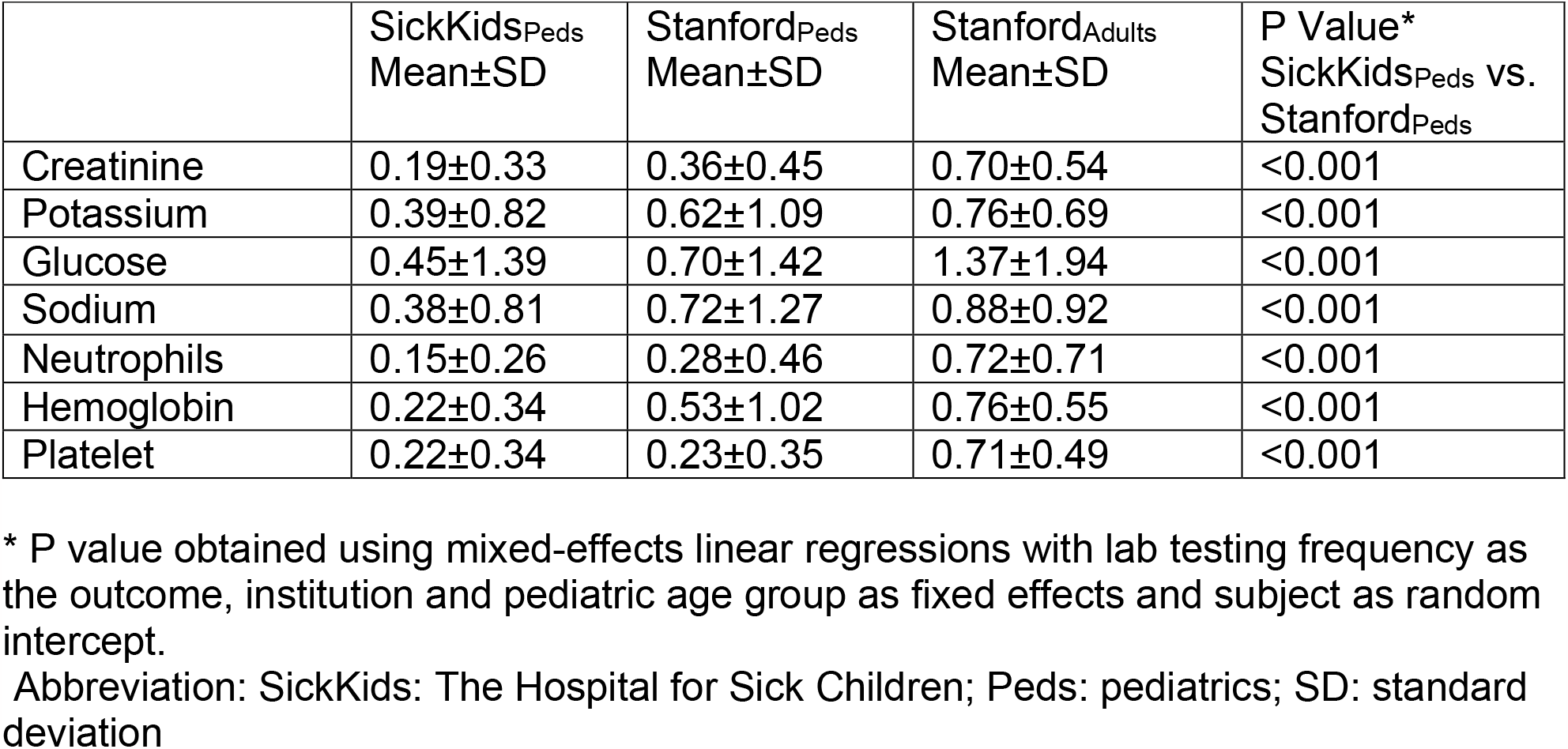
Average number of lab tests per inpatient day across all admissions by cohort.

|  | SickKids<sub>Peds</sub><br>Mean±SD | Stanford<sub>Peds</sub><br>Mean±SD | Stanford<sub>Adults</sub><br>Mean±SD | P Value*<br>SickKids<sub>Peds</sub> vs.<br>Stanford<sub>Peds</sub> |
| --- | --- | --- | --- | --- |
| Creatinine | 0.19±0.33 | 0.36±0.45 | 0.70±0.54 | <0.001 |
| Potassium | 0.39±0.82 | 0.62±1.09 | 0.76±0.69 | <0.001 |
| Glucose | 0.45±1.39 | 0.70±1.42 | 1.37±1.94 | <0.001 |
| Sodium | 0.38±0.81 | 0.72±1.27 | 0.88±0.92 | <0.001 |
| Neutrophils | 0.15±0.26 | 0.28±0.46 | 0.72±0.71 | <0.001 |
| Hemoglobin | 0.22±0.34 | 0.53±1.02 | 0.76±0.55 | <0.001 |
| Platelet | 0.22±0.34 | 0.23±0.35 | 0.71±0.49 | <0.001 |

\* P value obtained using mixed-effects linear regressions with lab testing frequency as the outcome, institution and pediatric age group as fixed effects and subject as random intercept.
Abbreviation: SickKids: The Hospital for Sick Children; Peds: pediatrics; SD: standard deviation

